# Blood transcriptomes of anti-SARS-CoV2 antibody positive healthy individuals with prior asymptomatic versus clinical infection

**DOI:** 10.1101/2021.04.19.21255748

**Authors:** Petros P. Sfikakis, Kleio-Maria Verrou, Ourania Tsitsilonis, Dimitrios Paraskevis, Efstathios Kastritis, Evi Lianidou, Paraskevi Moutsatsou, Evangelos Terpos, Ioannis Trougakos, Vasiliki Chini, Menelaos Manoloukos, Panagiotis Moulos, Georgios A. Pavlopoulos, George Kollias, Giannis Ampatziadis-Michailidis, Pantelis Hatzis, Meletios A Dimopoulos

## Abstract

Despite tremendous efforts by the international research community to understand the pathophysiology of SARS-CoV-2 infection, the reasons behind the clinical variability, ranging from asymptomatic infection to lethal disease, are still unclear. Existing inter-individual variations of the immune responses, due to environmental exposures and genetic factors, may be critical to the development or not of symptomatic disease after infection with SARS-CoV-2, and transcriptomic differences marking such responses may be observed even later, after convalescence. Herein, we performed genome-wide transcriptional whole-blood profiling to test the hypothesis that immune response-related gene signatures may differ between healthy individuals with prior entirely asymptomatic versus clinical SARS-CoV-2 infection, all of which developed an equally robust antibody response. Among 12.789 protein-coding genes analyzed, there were only six and nine genes with significantly decreased or increased expression, respectively, in those with prior asymptomatic infection (n=17, mean age 34 years) relatively to those with clinical infection (n=15, mean age 37 years). All six genes with decreased expression (*IFIT3, IFI44L, RSAD2, FOLR3, PI3, ALOX15*), are involved in innate immune response while the first two are interferon-induced proteins. Among genes with increased expression six are involved in immune response (*GZMH, CLEC1B, CLEC12A*), viral mRNA translation (*GCAT*), energy metabolism (*CACNA2D2*) and oxidative stress response (*ENC1*). Notably, 8/15 differentially expressed genes are regulated by interferons. Our results suggest that an intrinsically weaker expression of some innate immunity-related genes may be associated with an asymptomatic disease course in SARS-CoV-2 infection. Whether a certain gene signature predicts, or not, those who will develop a more efficient immune response upon exposure to SARS-CoV-2, with implications for prioritization for vaccination, warrant further study.

## Introduction

Since December 2019 the SARS-CoV-2 has spread throughout the world infecting dozens of millions of people and resulting in over 2.8 million deaths, as of April 2021. Although the case fatality rate in hospitalized patients may exceed 10% [1, 2], 35-50% of infected adults do not develop, perceive and report any clinical symptom [3, 4]. Asymptomatic infected persons may be responsible for viral transmission for more days than aware self-isolated cases, which may also explain, at least partially, the exponential increase in the number of infections globally [5, 6, 7]. Notably, we only know in retrospect who was indeed asymptomatic, since individuals without symptoms at the time of a positive molecular test should be followed for 14 days to determine the clinical picture, being “pre-symptomatic” if they develop symptoms later.

The proportion of asymptomatic individuals varies widely in viral infections. For example, a significant fraction of cytomegalovirus infections, similarly to SARS-CoV-2, are asymptomatic and unsuspected [8]. In contrast, an asymptomatic carrier state has not been documented for measles virus infection [9]. The reasons why certain individuals, including people living with HIV [10] or other immunodeficiencies [11], do not develop clinical symptoms during SARS-CoV-2 infection are essentially unknown [12]. So far, studies assessing the immune response in asymptomatic infection are few. In an elegant study, Long et al. showed that asymptomatic individuals presented with significantly longer duration of viral shedding compared to symptomatic patients, lower levels of IgG antibodies to SARS-CoV-2, and lower serum levels of 18/48 cytokines, including interferon-gamma levels, suggesting that asymptomatic individuals indeed displayed a weaker anti-virus-reactive immune response to SARS-CoV-2 [13].

While the role of genetics in determining immune and clinical response to the SARS-CoV-2 virus is currently under investigation [14], it is well established that individual human immune systems are highly variable [15]. Most of this inter-individual immune variation is explained by environmental exposures early in life [16] but genetic factors are clearly also involved. For example, a gene expression signature dominated by interferon-inducible genes in the blood is prominent in systemic lupus erythematosus [17], whereas interferon-α is increased not only in the serum of these patients but also in their healthy first-degree relatives [18] pointing to genetic influences on the interferon-mediated immune interactions.

Clearly, the most successful response against SARS-CoV-2 occurs in those individuals who, while remaining asymptomatic, develop a robust adaptive immune response. We have recently examined the humoral immune response to SARS-CoV-2 in members of the National and Kapodistrian University of Athens, Greece [19]. Overall, among 4.996 people the unweighted seroprevalence of SARS-CoV-2 antibodies was 1.58%, whereas 49% of the seropositive individuals denied having had any clinical symptom compatible with previous SARS-CoV-2 infection, which was also unsuspected for 33% of them. Interestingly, in our study, the mean levels of antibodies to both the nucleocapsid (N) protein and the receptor-binding-domain (RBD) of the spike (S) protein were comparable between asymptomatic and clinical infection cases and not associated with age or sex [4]. Others have also reported that IgG antibodies are commonly observed in both asymptomatic and clinical infections (85% versus 94% of patients, respectively) [20], whereas durable B cell-mediated immunity against SARS-CoV-2 after mild or severe disease occurs in most individuals [21].

Since variations in the strength and/or extent of the immune response may be critical for the clinical picture and progress after infection with SARS-CoV-2, existing inter-individual differences at the transcriptome level may be observed even later, after convalescence. Therefore, we performed 3’ mRNA next generation sequencing-based genome-wide transcriptional whole blood profiling to test the hypothesis that immune response-related genes are differentially expressed between healthy individuals who developed an equally robust antibody response following either an entirely asymptomatic or clinical SARS-CoV-2 infection.

## Methods

### Blood collection and anti-SARS-CoV-2 antibody testing

Blood samples were collected from members of the NKUA, Athens, Greece in June–November 2020. The protocol was approved by the Ethics and Bioethics Committee of the School of Medicine, NKUA (protocol #312/02-06-2020) and study participants provided written informed consent. All plasma samples were analyzed as previously described [4] using, a) the CE-IVD Roche Elecsys® Anti-SARS-CoV-2 test, an electrochemiluminescence immunoassay (ECLIA) for the detection of total antibodies (IgG, IgM, and IgA; pan-Ig) to SARS-CoV-2 N-protein (Roche Diagnostics GmbH, Mannheim, Germany), and b) the CE-IVD Roche Elecsys® Anti-SARS-CoV-2 S, an ECLIA for the quantitative determination of antibodies (including IgGs) to the SARS-CoV-2 S-protein RBD (Roche Diagnostics).

### 3’ mRNA sequencing, mapping, quality control, and quantifications

Total RNA was isolated from whole blood, stored in paxgene, using the ExtractionMonarch® Total RNA Miniprep Kit (NEB #T2010). Upon blood isolation, Monarch DNA/RNA Protection Reagent (supplied as a 2x concentrate) was added undiluted to an equal volume of blood. Addition of the protection reagent and the following RNA isolation was performed as described in the Kit’s manual for Total RNA Purification from Mammalian Whole Blood Samples.

After quantification on a NanoDrop ND-1000 (Thermofisher) and Bioanalyzer RNA 6000 Nano assay (Agilent), 140-300ng of total RNA from samples passing quality control were processed using the QuantSeq 3’ mRNA-Seq Library Prep Kit FWD (Lexogen, 015.96) for library preparation. Libraries were assessed for molarity and median library size using Bioanalyzer High Sensitivity DNA Analysis (Agilent, 5067-4626). After multiplexing and addition of 13% PhiX Control v3 (Illumina, FC-110-3001) as spike in, the NGS was performed on a NextSeq550 with NextSeq 500/550 High Output Kit v2.5 - 75 cycles (Illumina, 20024906).

The quality of FASTQ files was assessed using FastQC (version 0.11.9) [22]. The reads were mapped to the GRCh38 reference human genome using STAR, as part of a pipeline provided by Lexogen and BlueBee. After quality control, we obtained quantifications for ∼16.737 (12.789 protein coding) genes with more than five reads in more than 25% of the 17 asymptomatic and 15 clinical disease samples. Raw bam files, one for each sample, were summarized to a 3’UTR read counts table, using the Bioconductor package GenomicRanges [23], through metaseqR2 [24]. The gene counts table was normalized for inherent systematic or experimental biases (e.g., sequencing depth, gene length, GC content bias) using the Bioconductor package EDASeq [25]. For the downstream analysis, 12 hemoglobin *(HBQ1, HBG2, HBZ, HBA2, HBA1, HBM, HBZP1, HBE1, HBG1, HBD, HBBP1, HBB*) genes were removed from all samples.

### Blood immune cell subsets deconvolution

CIBERSORTx [26] was used to estimate the proportion of blood immune cell subsets for each individual. As a signature matrix, the LM22 signature matrix for 22 subsets obtained at the single cell level was used. The Mann-Whitney U test was applied in order to calculate the significance of the difference in distributions between the asymptomatic and clinical groups. Statistical significance and plotting was calculated with R.

### Differential gene expression

The resulting gene counts table was subjected to differential expression analysis (DEA) to compare individuals with a history of asymptomatic versus clinical (“symptomatic”) infection using the Bioconductor packages DESeq [27], edgeR [28], NOISeq[29], limma [30], NBPSeq [31], baySeq [32]. In order to combine the statistical significance from multiple algorithms and perform meta-analysis, the PANDORA weighted P-value across results method was applied through metaseqR2. Multidimensional scaling was also applied through metaseqR2. DAVID analysis [33] was performed for the increased and decreased genes, both for enriched Kyoto Encyclopedia of Genes and Genomes (KEGG) pathways and for biological processes [Gene Ontology (GO)]. For the prediction of enriched regulons in asymptomatic disease we used the TRRUST (v2) reference transcription factor (TF)–target interaction database [34] and enrichR [35] focusing on the ChEA prediction with the increased genes in asymptomatic disease as input. For the identification of interferon-regulated genes the inteferome database (v2) [36] was used.

## Results

### Whole blood transcriptional profiling and determination of immune cell subsets in seropositive asymptomatic versus clinical infection

As shown in **Table 1**, the two groups under study comprised 15 seropositive individuals with a history of clinical infection within 3-months (median) before sampling (9 men, mean age 34 years) and 17 seropositive individuals with entirely asymptomatic infection (11 men, mean age 37 years). Cases were considered asymptomatic in the absence of any symptoms since the onset of the pandemic, according to a detailed history obtained by a physician (absence of fever of any grade, fatigue, conjunctivitis/sweating coughs, headaches, respiratory distress/dyspnea, smell or taste loss, diarrhea). Clinical infections were in their majority of low to moderate severity.

**Table 1:** Demographics and antibody measurements.

| | Number of<br>Individuals (males) | Age, mean±SD<br>(range) | anti-SARS-CoV-2<br>N-protein Abs,<br>mean±SD (range) | anti- SARS-CoV-2<br>S-protein RBD<br>Abs, mean±SD<br>(range) |
| --- | --- | --- | --- | --- |
| **Clinical**<br>**Disease** | 15 (9) | 34±14 (18-57) | 38±39 (5-119) * | 179±255 (6-752) |
| **Asymptomatic**<br>**Disease** | 17 (11) | 37±17 (19-70) | 46±45 (1-166) ** | 122±131<br>(3-426)*** |

n=14\*, n=15\*\*, n=14\*\*\*

Age, sex distribution and levels of antibodies to both SARS-CoV-2 N-protein and the S-protein RBD were comparable between asymptomatic and clinical cases.

Whole blood-derived, 3’ mRNA next generation sequencing-based, genome-wide transcriptional profiling was performed and, overall, more than 386 million reads were generated. Genes with fewer than five counts in fewer than 25% of the samples were filtered out, resulting to 16.747 profiled genes, of which 12.789 were protein coding. Twelve hemoglobin genes (*HBQ1, HBG2, HBZ, HBA2, HBA1, HBM, HBZP1, HBE1, HBG1, HBD, HBBP1, HBB*) were removed. A multidimensional scaling (MDS) plot generated using all 16.737 expressed genes, in order to avoid gene-type biases, revealed no clear separation of the two sample groups (**Figure 1A**).

**Figure 1.**
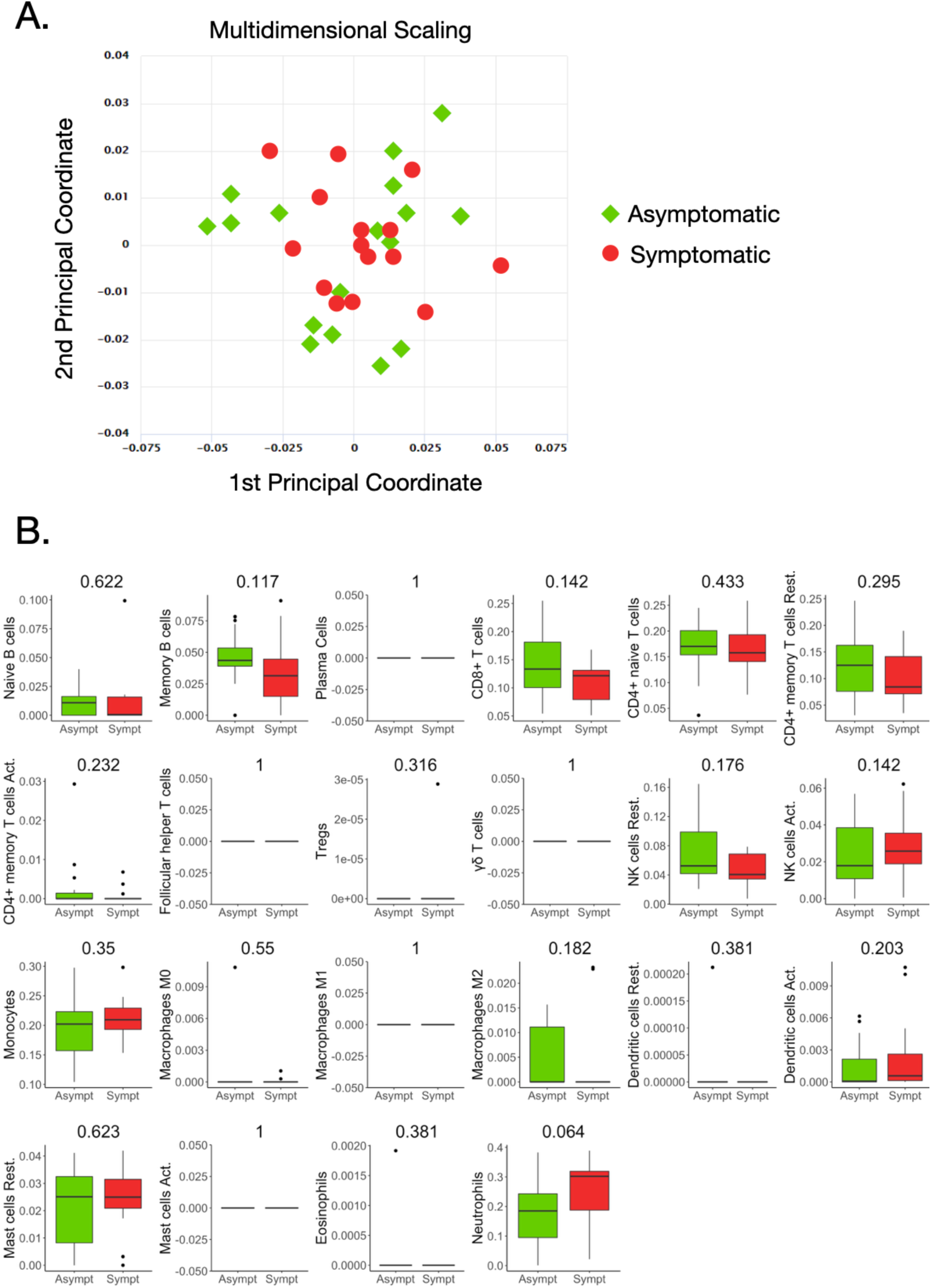
Whole blood transcriptional profiles and immune cell subsets in seropositive individuals with prior asymptomatic or clinical SARS-CoV-2 infection. (A) Dimensionality reduction of all samples: Multidimensional scaling of all samples from individuals with prior clinical (n=15) and asymptomatic (n=17) infection. Each dot corresponds to the sample of one individual. All expressed elements were used (16.747, out of which 12.799 were non-zero protein-coding genes), in order to avoid gene type biases. The smaller the distance between each sample pair, the greater the similarity of the gene expression profile of the samples. No separation of the two sample groups is revealed, reflecting their similarity. (B) Blood transcriptome deconvolution with CYBERSORTx in asymptomatic and clinical disease groups. For every cell type, the Mann-Whitney U test p-value comparing the two groups is displayed on top. No statistically significan differences (p-values < 0.05) were detected between the two groups.

The proportions of immune cell populations, namely, naive B cells, memory B cells, plasma cells, CD8+ T cells, naive CD4+ T cells, resting memory CD4+ T cells, activated memory CD4+ T cells, follicular helper T cells, regulatory T cells, gamma delta T cells, resting NK cells, activated NK cells, monocytes, M0 macrophages, M1 macrophages, M2 macrophages, resting dendritic cells, activated dendritic cells, resting mast cells, activated mast cells, eosinophils and neutrophils in the peripheral blood estimated by CIBERSORTx were also comparable between the two groups **(Figure 1B)**.

### Differentially expressed genes are associated with Innate Immunity and Interferon activity

Although the differential expression analysis of 12.789 protein coding genes did not reveal a distinct transcriptional profile between the two groups of healthy individuals, 24 genes were returned as differentially expressed (logFC=|1|, p<0.05) in a primary analysis (**S1 Figure**). Because the number of these genes was small, we repetitively applied the DEA pipeline, removing samples that were possible outliers in terms of expression of each differentially expressed gene. Therefore, genes that were repeatedly returned as significantly differentially expressed in those with prior asymptomatic infection relatively to those with clinical SARS-CoV-2 infection were characterized as differently expressed **(Figure 2)**. Brief description of the function of six and nine genes that were found significantly decreased (**S1 Table)** and increased (**S2 Table**), respectively, in prior asymptomatic versus clinical SARS-CoV-2 infection is shown in Supplemental Tables.

**Figure 2.**
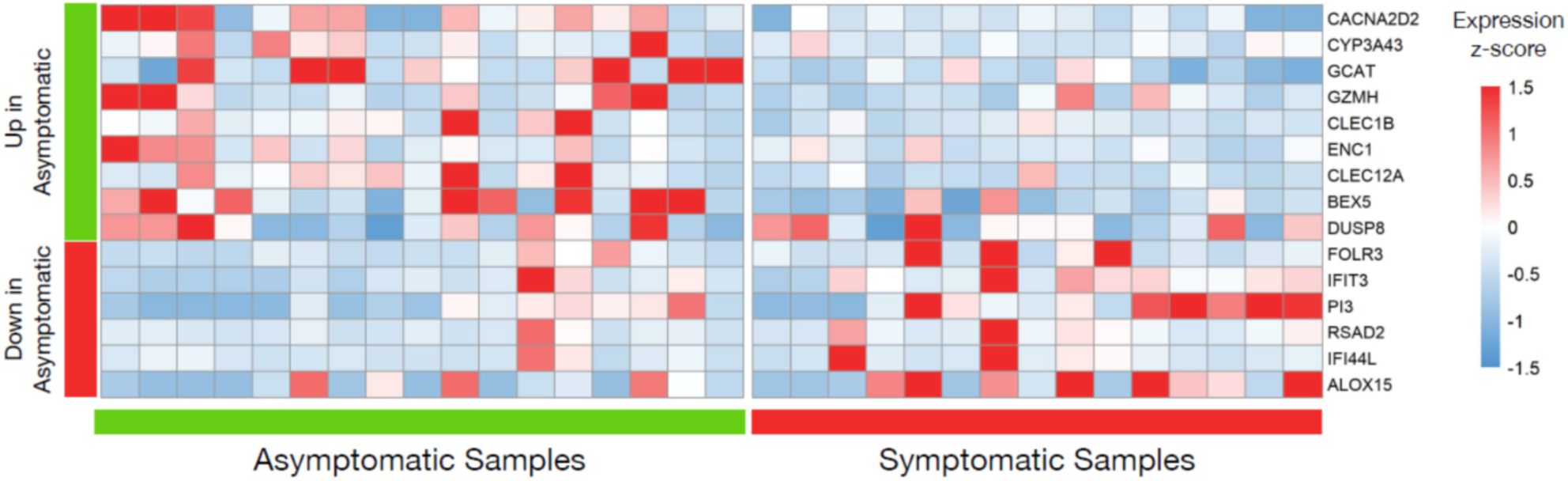
Differential gene expression analysis in seropositive individuals with prior asymptomatic or clinical SARS-CoV-2 infection. Heatmap of robustly differentially expressed genes (genes that were differentially expressed and highly expressed in three or more samples, logFC>|1|, p-value<0.05) in individuals with prior asymptomatic infection relatively to those with clinical (“symptomatic”) SARS-CoV-2 infection, with raw expression values being scaled. The values for all samples (17 asymptomatic on the left and 15 clinical on the right) is plotted. The first nine genes are increased in the Asymptomatic group, while the next six are decreased.

Notably, all six decreased genes in asymptomatic SARS-CoV-2 infection (*IFIT3, IFI44L, FOLR3, RSAD2, PI3, ALOX15*), are involved in innate immune responses [37-42] while the first two are interferon-inducible genes. Similarly, three increased genes (*GZMH, CLEC1B, CLEC12A*) are involved in innate immunity mechanisms [39,43,44], one (*GCAT*) in viral mRNA translation [45], one (*CACNA2D2*) in the integration of energy metabolism [46] and one (*ENC1*) in oxidative stress responses [47]. The expression patterns of these 15 genes across all samples are depicted in **Figure 2**.Enrichment analysis returned no statistically significant enriched KEGG or GO terms. Similarly, there were no common upstream transcriptional regulators revealed by transcription factor (TF)– target interaction databases for these genes.

Finally, the inteferome database which hosts genomic and transcriptomic data generated from cells or tissues treated with interferons was used for the 15 genes that were found to be differently expressed in asymptomatic versus clinical SARS-CoV-2 infections. Collectively, 8 out of 15 genes are regulated by interferons (*ENC1, FOLR3, IFIT3, PI3, RSAD2, IFI44L,CLEC12A, ALOX15*). Specifically, six genes are regulated by both type I and type II Interferons (*ENC1, FOLR3, IFIT3, PI3, RSAD2, IFI44L*), whereas the remaining two are targets of interferon type II only (*CLEC12A, ALOX15*) [36] **(Figure 3)**.

**Figure 3.**
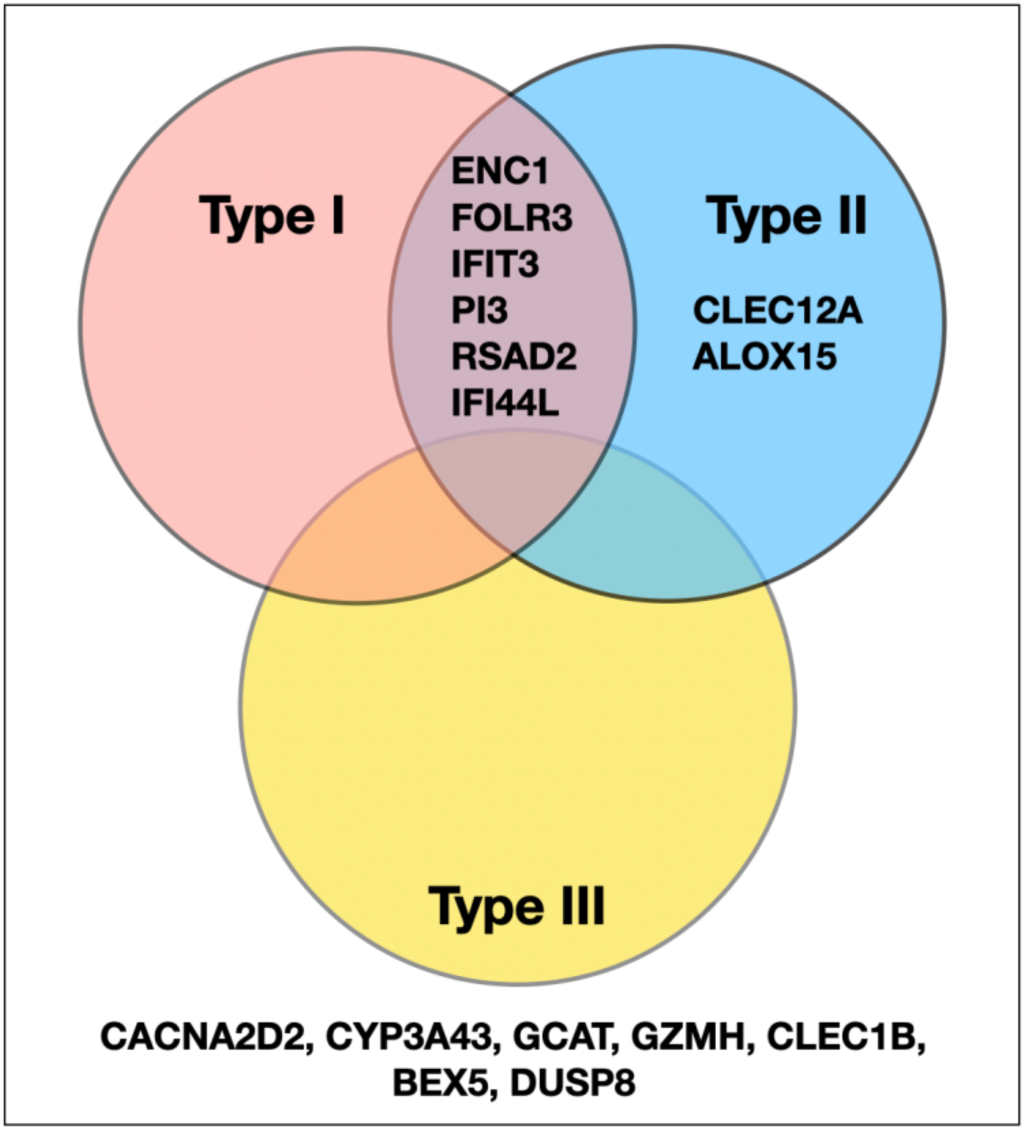
Venn diagram of differentially expressed genes in seropositive individuals with prior asymptomatic or clinical SARS-CoV-2 infection with respect to interferon activity. The genes characterized as differentially expressed in those with prior asymptomatic infection relatively to those with clinical SARS-CoV-2 infection were queried in the Interferome database; 8/15 were associated with interferon activity. Of those, six are regulated by both interferon type I and II, while two genes only by type II and none by type III; the remaining seven genes are not regulated by interferons.

## Discussion

Genome-wide transcriptome analyses studies using next generation sequencing technology in patients infected with SARS-CoV-2 provide evidence that transcriptome-wide changes may serve as predictors of morbidity and possibly of response to specific therapies [48]. In addition, transcriptomic analyses may provide mechanistic insights into certain complications associated with SARS-CoV-2 infection [49]. To our knowledge, this is the first whole blood genome-wide transcriptomic comparative analysis in healthy individuals who either recovered from a clinical SARS-CoV-2 infection or an entirely asymptomatic infection. In one previous study published so far in asymptomatic seropositive individuals infected during a super spreading event, the transcriptome in peripheral blood mononuclear cells was similar to that of seronegative highly exposed individuals from the same community. The putative time of infection of seropositive asymptomatic individuals was 4–6 weeks prior to sample collection, suggesting that the development of antibody response following viral exposure in asymptomatic cases is not necessarily associated with sustained alterations in the immune transcriptome [50].

Variations in innate immune system responses and cytokine networks could explain, at least in part, the wide heterogeneity in clinical presentation of SARS-CoV-2 infection [51]. The symptom that best reflects the potency of the immune response, namely fever, has been repeatedly shown to be a poor diagnostic marker in severe disease [52, 53]. Along these lines, it has been speculated that asymptomatic infection could be partly explained by the examples of altered innate immunity mechanisms operating in bats and pangolins. Despite carrying an enormous load of viral species, these animals display an apparent genetic resistance to coronavirus pathology [54]. For example, decomposition of many type I interferon genes [55] and partial loss of function in stimulator of interferon genes (STING) is observed in bats [56]. Regarding pangolins, recent findings suggest that these animals have lost interferon-ε [57] as well as interferon-induced with helicase C domain 1 (IFIH1), also known as IFIH1/MDA5 [58].

Our results provide evidence that among 12.789 genes, there were only 15 with significantly different expression when comparing healthy, relatively young individuals after convalescence from a previous entirely asymptomatic SARS-CoV-2 infection to those with a clinical infection history. While there were no apparent differences in cellular components and no specific immune deficiencies or co-morbidities to explain the different clinical presentation, the small number of differentially expressed genes is not surprising since the cohort comprised apparently healthy individuals. It should be highlighted that the transcriptome analysis was not performed at the time of active infection; thus certain potential differential responses may have been blunted during assessment after infection. This could also explain the lack of differentially expressed genes with >2-fold change in our primary analysis. However, such differential responses should be more robust at the time of infection and more genes and immune networks may be differentially expressed.

Among the six genes that were found with significantly decreased expression in previously asymptomatic cases relatively to clinical cases, and in line with our research hypothesis, all are involved in innate immune responses (**S1 Table)** and two of these genes *(IFIT3, IFI44L*) belong to the interferon-induced family of genes. Overall, 8 of the 15 differentially expressed genes in those with prior asymptomatic infection relatively to those with clinical SARS-CoV-2 infection can be found in datasets that include genes which have been implicated in interferon related signaling pathways in vitro [36].

As happens in all viral infections, type I interferon response plays a major protective role for the host because not only promotes viral clearance but also triggers a prolonged adaptive immune response [59]. Insights into the innate and adaptive immune responses to SARS-CoV-2 have been gained by many research efforts over the past year [49]. The innate immune responses that protect against disease and particularly the role of type I and III interferons have been addressed in numerous studies, mainly in patients with severe disease at the time of sampling. Important findings by Casanova and collaborators have shown that either neutralizing autoantibodies to type I interferons [59] or deleterious mutations in components involved in interferon induction or signaling [60] predispose patients to life-threatening infections. Along these lines, a highly impaired type I interferon response has been reported in patients with severe disease [61]. However, in contrast to these findings, increased levels of interferons and interferon-stimulated genes have been observed in severe and life-threatening infections in many other studies [63, 64, 65]. Indeed, increased interferon-alpha levels are a biomarker of mortality [66].

Moreover, the SARS-CoV-2 receptor ACE2, which is expressed in specific cell subsets across tissues is an interferon-stimulated gene in human airway epithelial cells [67], suggesting that a weaker individual interferon response may be protective. The latter may explain the low infection levels and morbidity in children [52, 53] who, relative to adults, display, in general lower interferon responses [67] and lower ACE2 expression [69]. Taken together, in individuals infected with SARS-CoV-2, interferon-mediated responses may be protective or detrimental depending on the timing and the stage of infection, in addition to other factors, including viral load, age, and co-morbidities [59, 70, 71].

To conclude, our results suggest that subtle differences in the expression levels of innate immunity-related genes, including lower expression of genes involved in interferon signaling, may be beneficial for the host upon SARS-CoV-2 infection. The current study attempts to fill the existing gap regarding the potential implication of certain pathways in the clinical phenotype of SARS-CoV-2 infection. The described association of a subtle immune response to SARS-CoV-2 with a lack of clinical symptoms needs further investigation, which hopefully will be performed in the near future by established consortia [14] or other groups. Whether a certain innate immunity signature predicts, or not, those who will develop a more successful immune response upon contact with SARS-CoV-2, with possible implications for prioritization of vaccination, warrant further study.

## Data Availability

Data will be made available upon peer-review in GEO.

## Acknowledgements

We thank Professor Argyrios Theofilopoulos for critical review of the manuscript. We acknowledge financial support of this work by the project pMedGR (MIS 5002802), funded by the Operational Programme “Competitiveness, Entrepreneurship and Innovation’ (NSRF 2014-2020) and co-financed by Greece and the European Union (European Regional Development Fund)”.

