## Supplementary material for "Blood transcriptomes of anti-SARS-CoV2 antibody positive healthy individuals with prior asymptomatic versus clinical infection": S1 Table

### Appendix

### S1 Table

Information on the function of genes with decreased expression in prior asymptomatic versus clinical SARS-CoV-2 infection

| Gene | Description and Biological Terms Related (if applicable) |
|---|---|
| **IFIT3** | Interferon Induced Protein With Tetratricopeptide Repeats 3<br>Pathways: Innate Immune System and Interferon gamma signaling.<br>Terms: identicalproteinbinding. |
| **IFI44L** | Interferon Induced Protein 44 Like<br>Diseases associated: Lymph Node Tuberculosis and Immunodeficiency 38 With Basal Ganglia Calcification. |
| **FOLR3** | Folate Receptor Gamma<br>Pathways: Innate Immune System and Endocytosis.<br>Terms: folicacidbinding. |
| **PI3** | Peptidase Inhibitor 3.<br>Pathways: Innate Immune System and Developmental Biology.<br>Terms: serine-type endopeptidase inhibitor activity and peptidase inhibitor activity. |
| **RSAD2** | Radical S-Adenosyl Methionine Domain Containing 2 (Viperin)<br>Pathways: Innate Immune System and Interferon gamma signaling.<br>Terms: self-association andiron-sulfurclusterbinding. |
| **ALOX15** | Arachidonate 15-Lipoxygenase<br>The enzyme acts on various polyunsaturated fatty acid substrates to generate various bioactive lipid mediators such as eicosanoids, hepoxilins, lipoxins, and other molecules. The encoded enzyme and its reaction products have been shown to regulate inflammation and immunity. [RefSeq, Aug 2017]<br>Pathways: Interleukin-4 and 13 signaling and Arachidonic acid metabolism.<br>Terms: iron ion binding and oxidoreductase activity, acting on single donors with incorporation of molecular oxygen, incorporation of two atoms of oxygen. |
