## Supplementary material for "Blood transcriptomes of anti-SARS-CoV2 antibody positive healthy individuals with prior asymptomatic versus clinical infection": S2 Table

Information on the function of genes with increased expression in prior asymptomatic versus clinical SARS-CoV-2 infection

| Gene | Description and Biological Terms Related (if applicable) |
|---|---|
| **DUSP8** | Dual Specificity Phosphatase 8<br>Pathways: Signaling by GPCR and MAPK signaling pathway<br>Terms: phosphatase activity and protein tyrosine/serine/threonine phosphatase activity |
| **CACNA2D2** | Calcium Voltage-Gated Channel Auxiliary Subunit Alpha2delta 2<br>Pathways: ERK Signaling and Activation of cAMP-Dependent PKA.<br>Terms: voltage-gated calcium channel activity and calcium channel regulator activity |
| **GCAT** | Glycine C-Acetyltransferase<br>Pathways: Glycine, serine and threonine metabolism, Viral mRNA Translation.<br>Terms: pyridoxalphosphatebinding and glycine C-acetyltransferaseactivity. |
| **GZMH** | GranezymeH<br>Reported to be constitutively expressed in the NK (natural killer) cells of the immune system and may play a role in the cytotoxic arm of the innate immune response by inducing target cell death and by directly cleaving substrates in pathogen-infected cells. [RefSeq, Nov 2015]<br>Pathways are Peptide hormone metabolism and Metabolism of proteins.<br>Terms: serine-type endopeptidase activity. |
| **CLEC12A**<br>**(MICL)** | C-lectine-like receptor<br>Pathways are Innate Immune System and C-type lectin receptor signaling pathway.<br>Terms: includecarbohydratebinding. |
| **CLEC1B**<br>**(CLEC2)** | C-Type Lectin Domain Family 1 Member B<br>Natural killer (NK) cells express multiple calcium-dependent (C-type) lectin-like receptors, such as CD94 and NKG2D, that interact with major histocompatibility complex class I molecules and either inhibit or activate cytotoxicity and cytokine secretion. CLEC2 is a C-type lectin-like receptor expressed in myeloid cells and NK cell (Colonna et al., 2000 [PubMed 10671229])<br>Pathways: Response to elevated platelet cytosolic Ca2+ and C-type lectin receptor signaling pathway.<br>Terms: transmembrane signaling receptor activity and carbohydrate binding. |
| **BEX5** | Brain expressed X-linked5, Nerve Growth Factor Receptor-Associated Protein 2 [Alvarez et al. 2005] |
| **ENC1** | Ectodermal-Neural Cortex 1<br>Pathways are WNT Signaling.<br>Terms: actinbinding. |
| **CYP3A43** | Cytochrome P450 Family 3 Subfamily A Member 43<br>Pathways are Paroxetine Pathway, Pharmacokinetics and Drug metabolism - cytochrome P450.<br>Terms: iron ion binding and oxidoreductase activity, acting on paired donors, with incorporation or reduction of molecular oxygen |
