## Supplementary figures and images for "Blood transcriptomes of anti-SARS-CoV2 antibody positive healthy individuals with prior asymptomatic versus clinical infection"

### S1 Figure

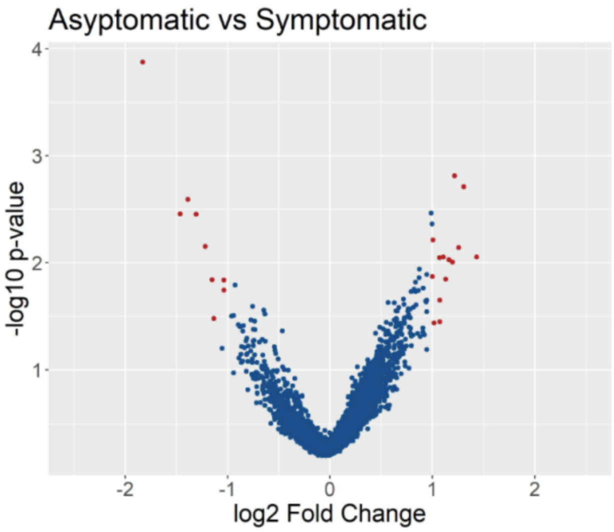

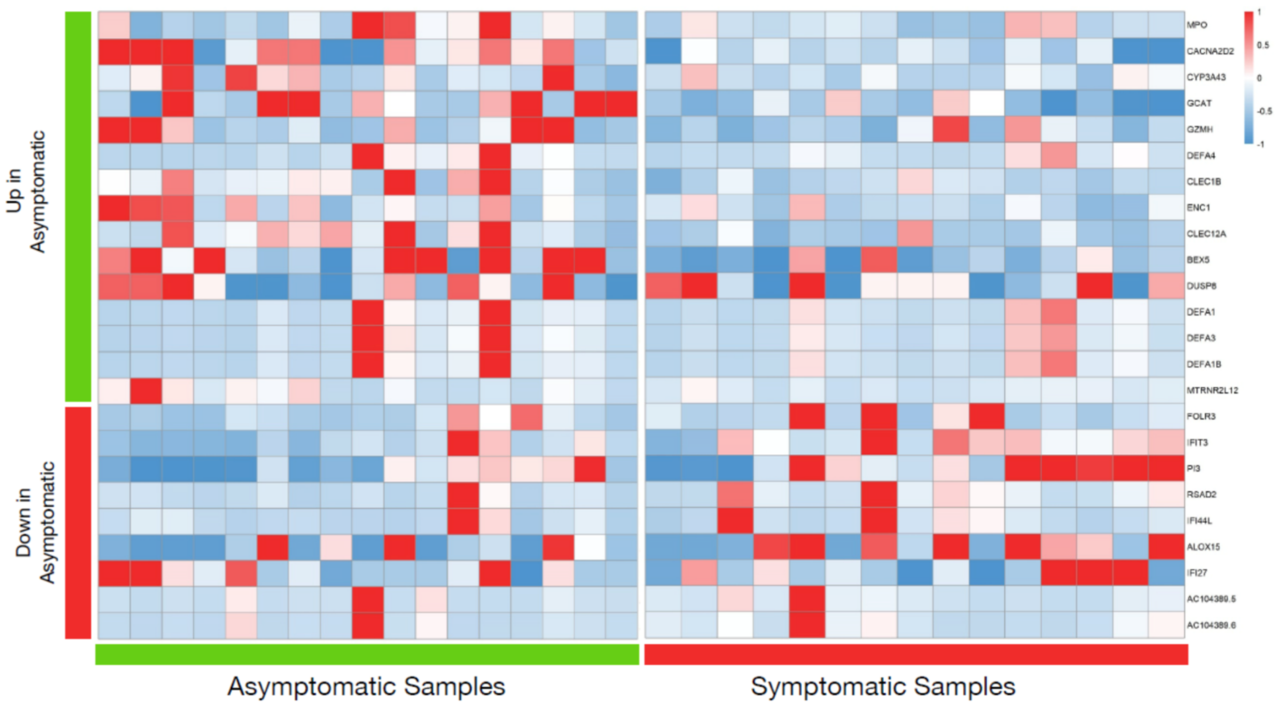
